# Deep Learning-based Multiclass Segmentation in Aneurysmal Subarachnoid Hemorrhage

**DOI:** 10.1101/2024.06.24.24309431

**Authors:** Julia Kiewitz, Orhun Utku Aydin, Adam Hilbert, Marie Gultom, Anouar Nouri, Ahmed A Khalil, Peter Vajkoczy, Satoru Tanioka, Fujimaro Ishida, Nora F. Dengler, Dietmar Frey

**Affiliations:** CLAIM - Charité Lab for AI in Medicine, Charité Universitätsmedizin Berlin, Corporate Member of Freie Universität Berlin, Humboldt-Universität zu Berlin and Berlin Institute of Health, Charitéplatz 1, 10117, Berlin, Germany; Department of Neurosurgery, Charité Universitätsmedizin Berlin, Corporate Member of Freie Universität Berlin, Humboldt-Universität Zu Berlin and Berlin Institute of Health, Charitéplatz 1, 10117, Berlin, Germany; Centre for Stroke Research Berlin, Charité Universitätsmedizin Berlin, Berlin, Germany; Department of Neurosurgery, Mie Chuo Medical Center, 2158-5 Myojin-cho, Hisai, Tsu, Mie 5141101, Japan; Faculty of Health Sciences Brandenburg, Medical School Theodor Fontane, Campus Bad Saarow, Germany; Department of Neurosurgery, HELIOS Hospital Bad Saarow, Germany

**Author notes:** **Corresponding Author Details** PD Dr. med. Dietmar Frey Charité Lab for AI in Medicine Charité Universitätsmedizin Berlin Charitéplatz 1 10117 Berlin Germany.

**Keywords:** subarachnoid hemorrhage, deep learning, multiclass segmentation, interrater reliability, outcome prediction

## Abstract

**Introduction:** Aneurysmal subarachnoid hemorrhage (aSAH) is a life-threatening condition with a significant variability in patients’ outcomes. Radiographic scores used to assess the extent of SAH or other potentially outcome-relevant pathologies are limited by interrater variability and do not utilize all available information from the imaging. Image segmentation plays an important role in extracting relevant information from images by enabling precise identification and delineation of objects or regions of interest. Thus, segmentation offers the potential for automatization of score assessments and downstream outcome prediction using precise volumetric information. Our study aims to develop a deep learning model that enables automated multiclass segmentation of structures and pathologies relevant for aSAH outcome prediction.

**Methods:** Out of 408 patients treated with aSAH in the department of Neurosurgery at Charité University Hospital in Berlin from 2009 to 2015, a subset of 73 representative CT scans were included in our retrospective study. All non-contrast CT scans (NCCT) were manually segmented to create a ground truth. For the multiclass segmentation task we determined six different target classes: basal and cortical SAH, intraventricular hemorrhage (IVH), ventricles, intracerebral hemorrhage (ICH), aneurysms and subdural hematoma (SDH). An additional hemorrhage class was created by merging the individual hemorrhage classes. The set of 73 NCCT was splitted into three stratified sets: training set (43 patients), validation set (10 patients) and test set (20 patients). We used the nnUnet deep learning based biomedical image segmentation tool and implemented 2d and 3d configurations. Additionally, we performed an interrater reliability analysis for multiclass segmentation and assessed the generalizability of the model on an external dataset of primary ICH patients (n=104). Segmentation performance was evaluated using: median Dice coefficient, volumetric similarity and sensitivity. Additionally, a global Dice coefficient was calculated by considering all patients in the test set to be one single concatenated image.

**Results:** The nnUnet-based segmentation model demonstrated performance closely matching the interrater reliability observed between two senior human raters for the SAH, ventricles, ICH classes and overall hemorrhage segmentation. For the hemorrhage segmentation a global Dice coefficient of 0.730 was achieved by the 3d model and a global Dice coefficient of 0.736 was achieved by the 2d model. The global Dice coefficient of the SAH class was 0.686 for both of the nnUnet models; ICH: 0.743 (3d model), 0.765 (2d model); ventricles: 0.875 (3d model), 0.872 (2d model). In the IVH, aneurysm and SDH classes the nnUnet models performance differed the most from the human level performance. Overall, the interrater reliability of the SAH class was observed to be lower than in other classes. In the external test set a global Dice coefficient of 0.838 for the hemorrhage segmentation was achieved.

**Conclusion:** Deep learning enables automated multiclass segmentation of aSAH-related pathologies and achieves performance approaching that of a human rater. This enables automatized volumetries of pathologies identified on admission CTs in aSAH patients potentially leading to imaging biomarkers for improved aSAH outcome prediction.

## Introduction

Aneurysmal subarachnoid hemorrhage (aSAH) is a severe subtype of stroke with an incidence of 8/100,000 person-years leading to a potential loss of many years of productive life (1,2). Patient outcomes following aSAH show significant variability; while aSAH is lethal for one-third of patients, survivors may become permanently dependent on nursing care due to residual cognitive and functional impairment (3). In aSAH several clinical and imaging variables were shown to predict outcome. The initial imaging performed for diagnosing SAH is non-contrast CT imaging (NCCT) and delivers important information on the presence, volume, location and radiological appearance of the hemorrhage (4).

Different radiographic scores are used to assess the extent of the subarachnoid hemorrhage and highlight different aspects of SAH. For example, SEBES (subarachnoid hemorrhage early brain edema score) predicts delayed cerebral ischemia (DCI) and unfavorable outcomes (5), the Graeb Score focuses on intraventricular bleeding (6) and the Hijdra Sum Score includes a grading system based on the amount of subarachnoid blood in different localizations of the brain (7). A different common scoring system is the Barrow Neurological Institute (BNI) scoring system which estimates the risk of vasospasm depending on the radiographic extent of SAH in NCCT via measuring the thickness of the hemorrhage and distinguishing five different severity levels (8). Out of all radiographic scores the modified Fisher Scale (9) is predominantly used in clinical practice due to its practicality. It quantifies the amount of subarachnoid blood to predict cerebral vasospasm, a condition leading to poor outcome and high mortality.

A main limitation of radiographic scores is their subjective nature and considerable interrater variability, which may limit their predictive value and clinical utility. For instance, the Fisher Scale does not include an exact definition of the various scoring criteria – thick subarachnoid clot or presence of intracerebral and intraventricular hemorrhage – determining the different severity levels (10,11). The vague definition of imaging findings further reduces the interrater agreement which can be measured by weighted kappa score k_w_ reported for various scores (k_w_ for the Fisher Scale depending on different studies = 0.66/0.53/0.45) (10–14). The radiographic scores only focus on specific aspects of the images like blood thickness, location, or volume, without providing a comprehensive assessment of available information in the medical images.

Recently, several studies have proposed quantifying the SAH volume using automated segmentation methods in NCCT (15–18). There are different methods for binary segmentation of subarachnoid hemorrhage including threshold-based (16), semi-automated (17), and deep-learning-based methods (15,19) not only quantifying the SAH-volume but also trying to predict parameters that are relevant to the patient’s outcome – e.g. the prediction of vasospasm risk (17). Whereas a manual segmentation provided by an experienced human rater is considered the gold standard (17), this has several limitations including the high time effort and relatively low interrater reliability (15,16). Deep learning based methods have the potential to provide a more objective assessment of hemorrhage volumes in an automated way. Therefore, hemorrhage segmentation is addressed as part of segmentation challenges such as the INSTANCE challenge 2022 and the upcoming MICCAI 2024 challenge MBH-Seg for segmentation of intracranial hemorrhage (20–22). One limitation of recently proposed models is that they only provide binary labels for intracranial hemorrhage subtypes or do not explicitly consider various co-occurring pathologies and relevant anatomical changes such as enlarged or shifted ventricles. Hence the automated segmentation can only differentiate between healthy brain tissue and hemorrhage and can not classify the different types of hemorrhage. The distinction of the hemorrhage types is especially important because the presence of co-occurring pathologies – like ICH together with SAH – significantly worsens the outcome of aSAH patients (23,24). One of the first studies that assess multiclass segmentation of intracranial hemorrhage in patients after traumatic brain injury was published as a preprint in 2023 by Wu et al. (25). In contrast to our study, Wu et al. only assessed pathologies e.g. different types of intracranial hemorrhages and did not consider anatomical structures which, when altered, may also be of prognostic value.

Our study aims to develop a multiclass deep learning-based segmentation model tailored to aneurysmal subarachnoid hemorrhage that includes outcome-relevant structures and pathologies for rapid and accurate volume segmentation. The proposed method segments 6 different classes: basal and cortical SAH, intraventricular hemorrhage (IVH), ventricles, intracerebral hemorrhage (ICH), aneurysms visible on NCCT and subdural hematoma (SDH). Moreover, we assess expert interrater agreement and perform external validation of our model on patients with a different main pathology (primary ICH). Our model is made available open source with pre-trained weights to facilitate the extraction of outcome-related pathologies from NCCT images of aSAH patients for further research.

## Materials and Methods

### Patients

Out of 408 aSAH patients treated in the Department of Neurosurgery at Charité University Hospital in Berlin from 2009 to 2015, 73 patients were randomly selected and retrospectively included. The selection of a limited subset of 73 patients meeting the inclusion criteria was due to time constraints regarding manual segmentation (Figure 1). Inclusion criteria were aSAH; CT scan at admission; CT scan slice thickness between 4 to 6 mm; and patient’s age 18 or older. aSAH was diagnosed through non-contrast computed tomography (NCCT) scans, and if NCCT was negative, lumbar puncture was performed to assess bilirubin or other blood degradation products. 69 out of 73 patients had visible hemorrhage in NCCT and 4 patients were diagnosed with SAH by lumbar puncture because NCCT was negative.

**Figure 1:**
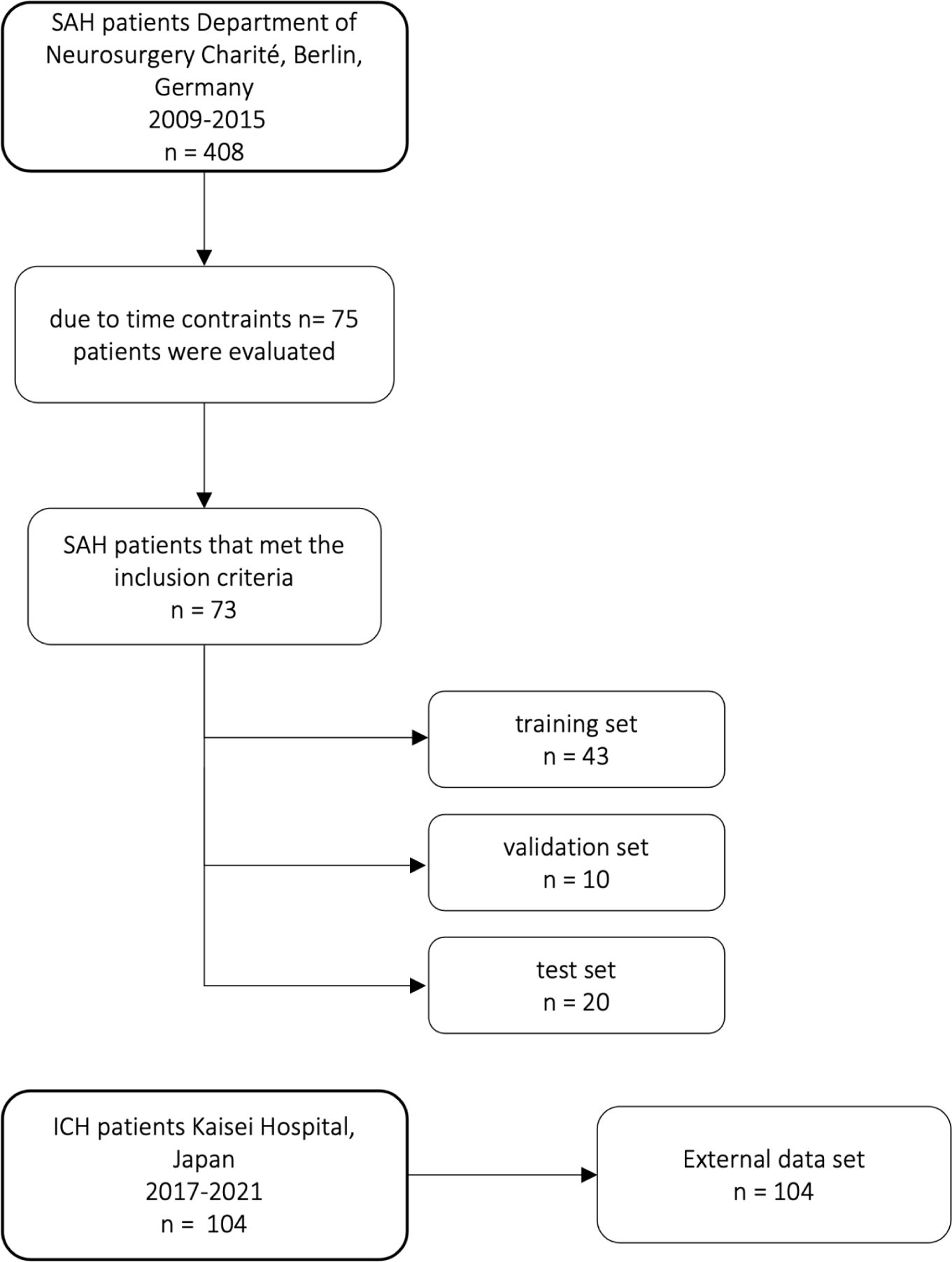
Flow chart of patients included in our study from the Department of Neurosurgery Charité University Hospital and distribution of training, validation, and test set and external data set from Kaisei Hospital, Japan.

To test for generalizability, we validated our final monocentric model on external data. For the external validation, we analyzed NCCT images of 104 patients presenting with the primary diagnosis of ICH at Kaisei Hospital, Japan.

Ethics approval was granted by the local authorities of Charité University Hospital (EA1/291/14) and Kaisei Hospital (2020-05) respectively.

### Image processing

The NCCT images were converted from DICOM (Digital imaging and communications in medicine) to NIfTI (Neuroimaging informatics technology initiative) file format using the dicom2niix command line tool in nipype (26). No gantry tilt correction was applied. Preprocessing of images, including reshaping, and resampling was performed by the default configuration of the nnUnet framework. All CT scans had a slice thickness of 4 to 6 mm (median slice thickness 5.0002 mm) with 24 to 32 slices (median number of slices: 28). The median voxel spacing after preprocessing was 5x0.44x0.44 mm and the median shape of the volumes were 512x512x28. For the external dataset the images were preprocessed manually to have uniform slice thickness of 2 mm with a shape of 512x512x80 using 3D slicer (27).

### Dataset and Labeling

73 head NCCT scans that met the inclusion criteria were manually segmented with ITK-Snap, an open-source 3d medical image analysis software (28) (ITK-SNAP Version 3.6.0). For the segmentation, six different target classes were determined: basal and cortical SAH, IVH, ventricles, ICH, aneurysms and SDH. All 73 CT scans were first manually segmented by a junior rater (JK) then checked and corrected by a senior rater (ND, senior physician neurosurgeon). The raters, except for JK, had no access to any other information about the patients besides the NCCT image. The manual segmentation of one CT scan took between 3 to 6 hours depending on the complexity and volume of the hemorrhages.

For the deep learning models, 73 CT scans were split into three sets: training set (n=43), validation set (n=10), and test set (n=20). The split was stratified by balancing the number of our six classes and the occurrence of metal artifacts and shunts in each set to avoid biases in the training of the nnUnet models (Table 3). Table 3 shows the distribution of the classes in the different sets.

For quality assurance, we calculated the interrater agreement. Another junior rater (MG) independently segmented the test set (20 out of the 73 CT scans) and another senior rater (AK, neuroradiology resident) checked and approved the segmentations to create a comparison group. In 4 patients we were only able to segment the ventricles as there was no visible hemorrhage in the NCCT, 2 of these patients belonged to the training set, one belonged to the validation set and one patient belonged to the test set.

Due to the time effort of multiclass segmentation of 3 to 6 hours per NCCT, we validated the results using an already-available binary hemorrhage segmentation dataset (hemorrhage or no hemorrhage) for the external validation of our models. The dataset was labeled by ST (senior physician neurosurgeon) using the software 3D slicer (27).

For the evaluation, the outputs of our multiclass 2d model were merged to create a single hemorrhage class.

### Deep Learning segmentation

nnUnet is a state-of-the-art biomedical image segmentation tool with automated configuration, including preprocessing, network architecture, training, and post-processing which gives the framework high flexibility (29). We also tested BRAVENET, which is a multiscale 3-D convolutional neural network (CNN) model initially developed on a dataset of patients with cerebrovascular diseases. For brain artery segmentation tasks, the BRAVENET architecture showed a superior performance compared to a standard Unet (30).

### Training

The nnUnet framework was trained in two configurations: 1) 3d and 2) 2d using axial slices. The models were trained for 1000 epochs. The best-performing model on the validation set after 1000 epochs was selected for evaluation on the test set. Model training was performed on an RTX Titan GPU with 24GB of VRAM, while the GPU VRAM argument was set to 8GB (default) for the training. The selected patch sizes, hyperparameters, and model architecture details can be found in Table 1. We used the 2d model for compatibility with various slice thickness values and across datasets and patients.

**Table 1:**
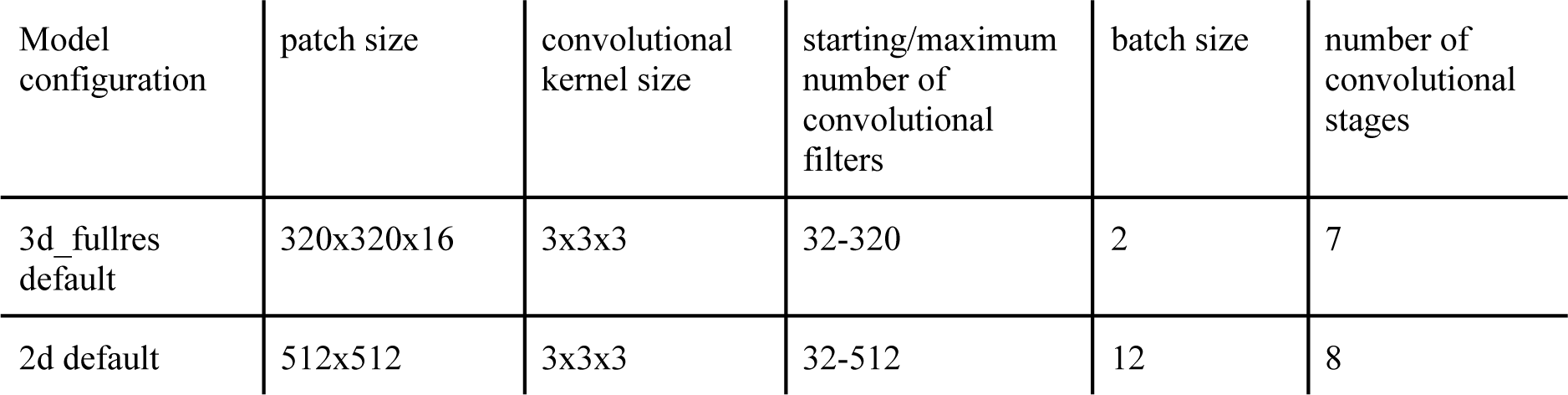
Model hyperparameters.

### Evaluation

The segmentation performance was evaluated using the EvaluateSegmentation tool (31). The quantitative evaluation of performance on each class was performed separately, and additionally a general hemorrhage group was created where all hemorrhage classes were merged to test the general ability of our model to detect blood in NCCT. To assess the performance, different metrics were reported: the median Dice coefficient, calculated on individual patients, the global Dice coefficient, by considering all patients in the test set to be one single concatenated image, volumetric similarity and sensitivity (Table 4). Classes where the model segmented less than 20 voxels were excluded from metric calculations.

The global Dice coefficient was calculated to provide a general overview of the model’s performance on the whole dataset (Equation 1), since in patients with smaller SAHs even small errors in segmentations can easily lead to loss of overlap and impact mean Dice values. The Dice coefficient was calculated according to following formula:

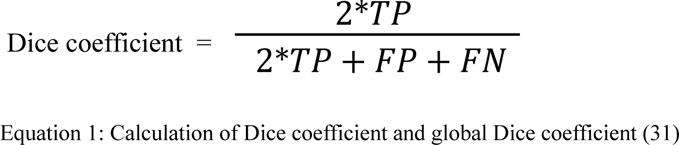

where TP (True Positives), FP (False Positives) and FN (False Negatives) are used from each individual patient. For calculating the global Dice coefficient the same Equation applies as the FP, TP and FN voxels across the entire test set were used.

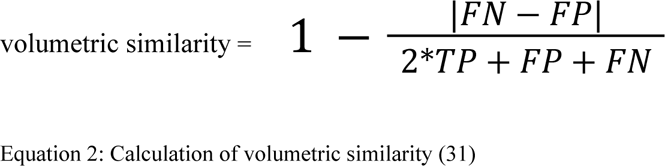

While the global Dice coefficient provides valuable insight into the spatial overlap between segmented and ground truth regions, we also calculated the volumetric similarity (Equation 2). Volumetric similarity directly emphasizes the portion of the segmented volume in relation to the reference volume and is an important measure for outcome prediction (31). Detailed descriptions and formulas of the metrics can be found in the study by Taha et. al. (31). Bland Altman Plots were created to show the volumetric similarity and bias of the models for the SAH class for the 3d and 2d nnUnet model and Rater 2 (Figure 4a) and the merged hemorrhage class (Appendix: Figure 4b). We also report the cases where the different models and the raters agreed or disagreed on the presence of a certain class.

## Results

We assessed 73 head CT scans from patients with aSAH at admission. Median age was 48 years [min;max = 22;92 years] and 63% of the patients were female (Table 2). Out of 73 CT scans, there were 71 with basal and/or cortical SAH, 42 with IVH, 25 with ICH, 9 with visible aneurysms, and 5 with SDH (Table 3). Out of 73 patients, 28 patients had image artifacts or additional treatment material: 4 had an intraventricular shunt, 19 had movement artifacts in the form of blur and 5 had metal artifacts due to previous aSAH therapy in the form of aneurysmal clipping or coiling.

**Table 2:** Patient characteristics.

|  | Training set =<br>43 | Validation set =<br>10 | Test set = 20 | n = 73 | External data<br>set = 104 |
| --- | --- | --- | --- | --- | --- |
| **sex** |  |  |  |  |  |
| male (%) | 17 (40%) | 0 (0%) | 10 (50%) | 27 (37%) | 56 (54%) |
| female (%) | 26 (60%) | 10 (100%) | 10 (50%) | 46 (63%) | 48 (46%) |
| **age** |  |  |  |  |  |
| mean age | 50 | 62 | 49 | 52 | 70 |
| median age | 46 (IQR = 19) | 62 (IQR = 24) | 49 (IQR = 12) | 48 (IQR = 17) | 71 (IQR = 23) |
| **Glasgow Coma Scale** |  |  |  |  |  |
| GCS < 9 | 25 | 5 | 7 | 37 | 15 |

**Table 3:**
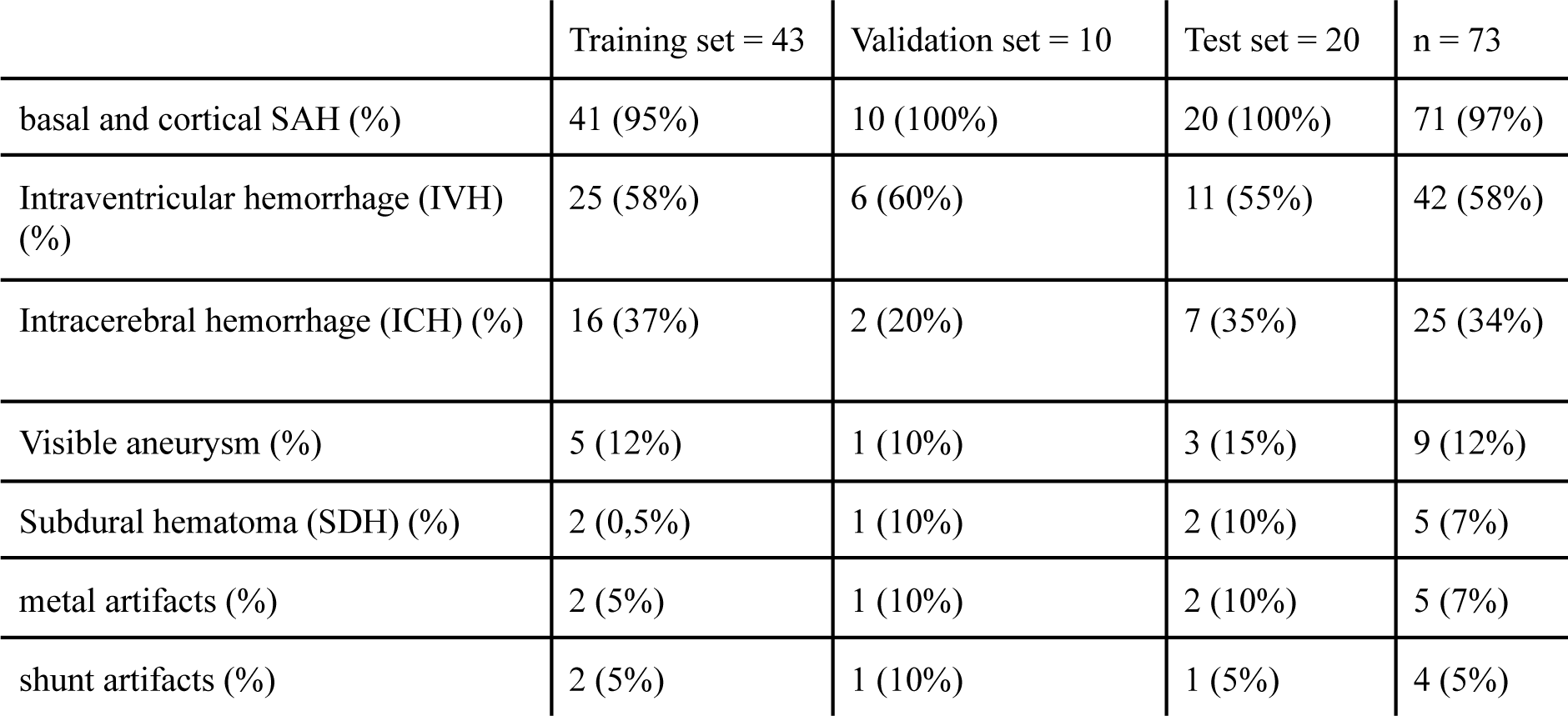
Distribution of different classes of the main data in training, validation and test set.

|  | Training set = 43 | Validation set = 10 | Test set = 20 | n = 73 |
| --- | --- | --- | --- | --- |
| basal and cortical SAH (%) | 41 (95%) | 10 (100%) | 20 (100%) | 71 (97%) |
| Intraventricular hemorrhage (IVH) (%) | 25 (58%) | 6 (60%) | 11 (55%) | 42 (58%) |
| Intracerebral hemorrhage (ICH) (%) | 16 (37%) | 2 (20%) | 7 (35%) | 25 (34%) |
| Visible aneurysm (%) | 5 (12%) | 1 (10%) | 3 (15%) | 9 (12%) |
| Subdural hematoma (SDH) (%) | 2 (0,5%) | 1 (10%) | 2 (10%) | 5 (7%) |
| metal artifacts (%) | 2 (5%) | 1 (10%) | 2 (10%) | 5 (7%) |
| shunt artifacts (%) | 2 (5%) | 1 (10%) | 1 (5%) | 4 (5%) |

### nnUNet segmentation results

In general, the nnUnet models achieved human-level performance for the SAH, the ICH, the ventricle and the hemorrhage segmentation class. For other classes such as the IVH, aneurysm, and the SDH class, the automated segmentation performance was lower than human-level performance (Table 4). Overall the 3d and 2d models performed similarly. The 3d model achieved a global Dice coefficient in the hemorrhage class of 0.730 and the 2d model achieved a global Dice coefficient of 0.736 (Table 4). For the SAH class, the 3d and 2d models achieved the same result with a global Dice coefficient of 0.686. The models had a slightly better performance for the ICH and IVH classes with a global Dice coefficient of 0.750 (IVH 3d and 2d model) and 0.743 (ICH 3d model) respectively 0.765 (ICH 2d model). The segmentation performance of the aneurysm class varied the most (3d model: global Dice coefficient = 0.037; 2d model: global Dice coefficient = 0.366). The results of the automatic segmentation of the 3d and 2d models of the SDH class were similar to the segmentation results of the IVH and ICH classes. The ventricle class segmentation achieved the highest global Dice coefficient of 0.875/0.872 (3d model/2d model).

**Table 4:**
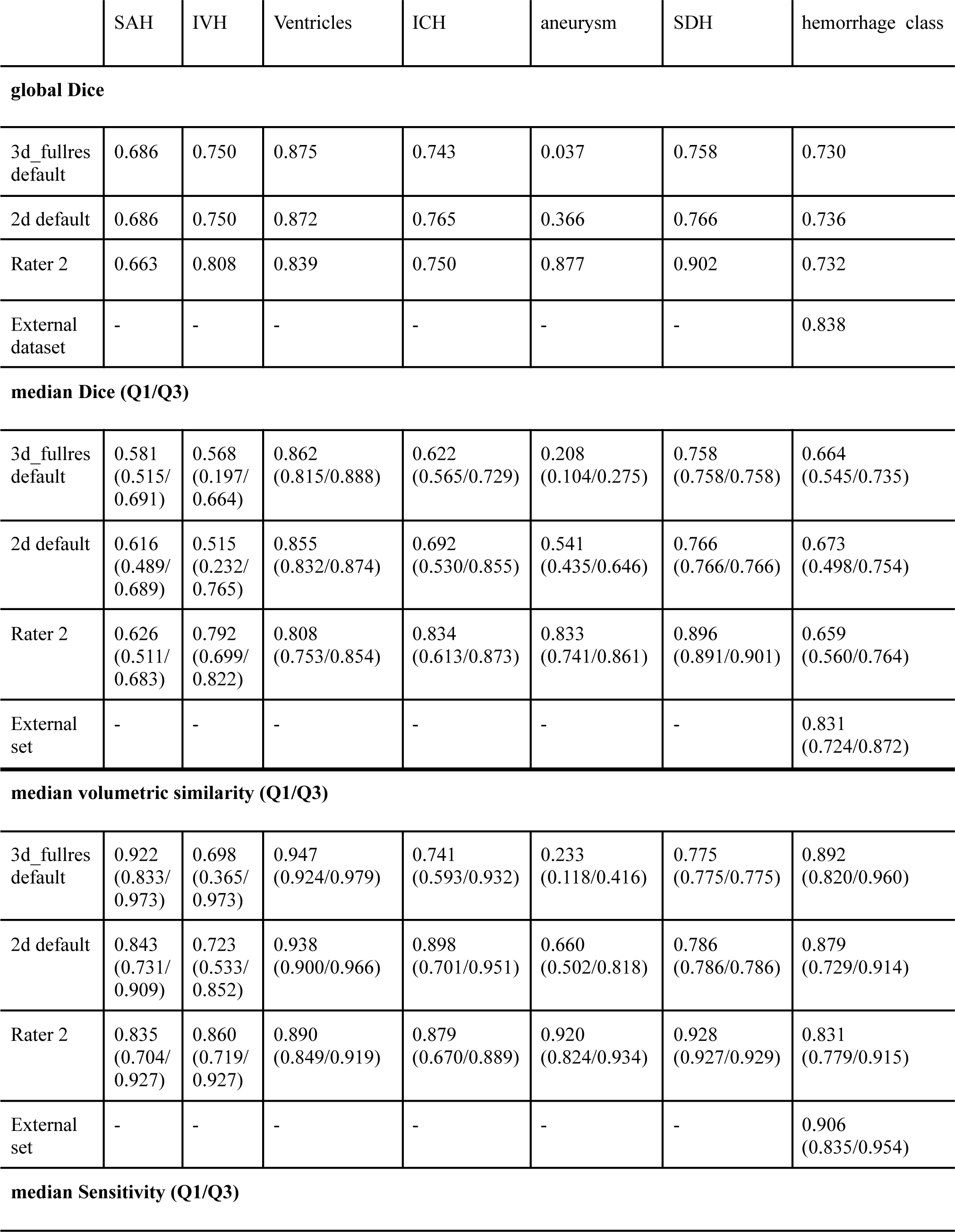
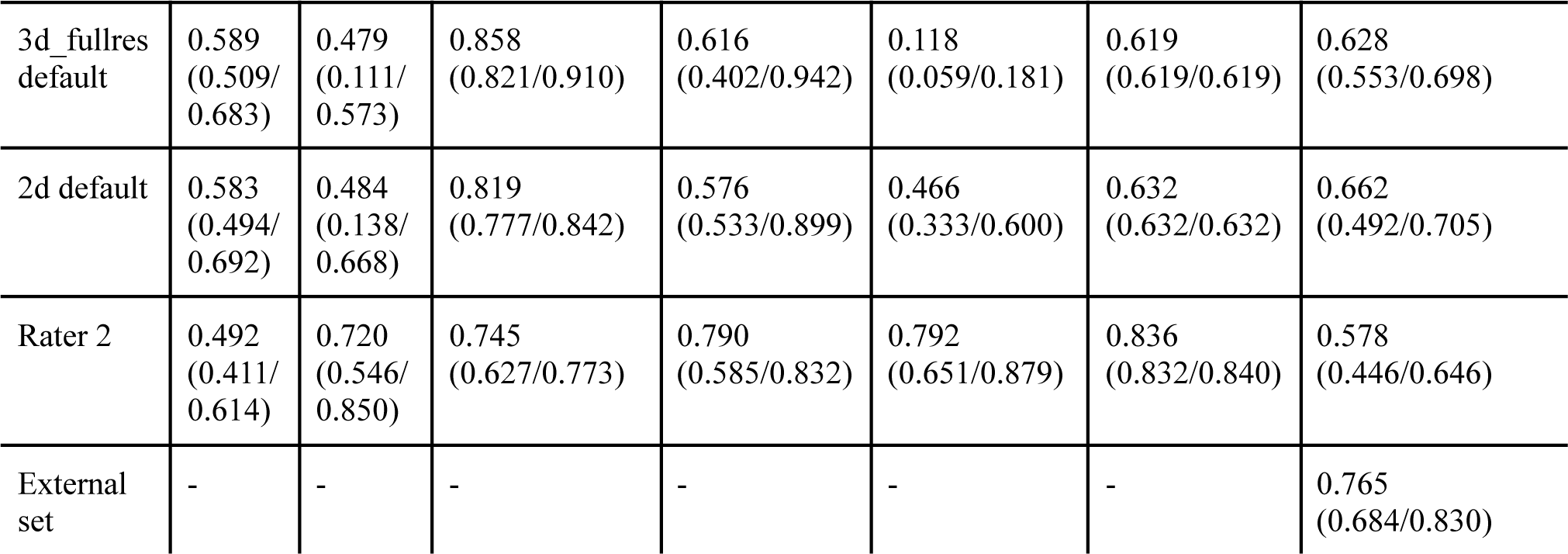
Comparison of different metrics for different labels for 3d model, 2d model, Rater 2 and the external dataset. Values were only calculated when there was an agreement on the presence of one class.

**global Dice**
|  |  |  |  |  |  |  |  |
| --- | --- | --- | --- | --- | --- | --- | --- |
| 3d_fullres default | 0.686 | 0.750 | 0.875 | 0.743 | 0.037 | 0.758 | 0.730 |
| 2d default | 0.686 | 0.750 | 0.872 | 0.765 | 0.366 | 0.766 | 0.736 |
| Rater 2 | 0.663 | 0.808 | 0.839 | 0.750 | 0.877 | 0.902 | 0.732 |
| External dataset | - | - | - | - | - | - | 0.838 |
| **median Dice (Q1/Q3)** |  |  |  |  |  |  |  |
| 3d_fullres default | 0.581<br>(0.515/<br>0.691) | 0.568<br>(0.197/<br>0.664) | 0.862<br>(0.815/0.888) | 0.622<br>(0.565/0.729) | 0.208<br>(0.104/0.275) | 0.758<br>(0.758/0.758) | 0.664<br>(0.545/0.735) |
| 2d default | 0.616<br>(0.489/<br>0.689) | 0.515<br>(0.232/<br>0.765) | 0.855<br>(0.832/0.874) | 0.692<br>(0.530/0.855) | 0.541<br>(0.435/0.646) | 0.766<br>(0.766/0.766) | 0.673<br>(0.498/0.754) |
| Rater 2 | 0.626<br>(0.511/<br>0.683) | 0.792<br>(0.699/<br>0.822) | 0.808<br>(0.753/0.854) | 0.834<br>(0.613/0.873) | 0.833<br>(0.741/0.861) | 0.896<br>(0.891/0.901) | 0.659<br>(0.560/0.764) |
| External set | - | - | - | - | - | - | 0.831<br>(0.724/0.872) |
| **median volumetric similarity (Q1/Q3)** |  |  |  |  |  |  |  |
| 3d_fullres default | 0.922<br>(0.833/<br>0.973) | 0.698<br>(0.365/<br>0.973) | 0.947<br>(0.924/0.979) | 0.741<br>(0.593/0.932) | 0.233<br>(0.118/0.416) | 0.775<br>(0.775/0.775) | 0.892<br>(0.820/0.960) |
| 2d default | 0.843<br>(0.731/<br>0.909) | 0.723<br>(0.533/<br>0.852) | 0.938<br>(0.900/0.966) | 0.898<br>(0.701/0.951) | 0.660<br>(0.502/0.818) | 0.786<br>(0.786/0.786) | 0.879<br>(0.729/0.914) |
| Rater 2 | 0.835<br>(0.704/<br>0.927) | 0.860<br>(0.719/<br>0.927) | 0.890<br>(0.849/0.919) | 0.879<br>(0.670/0.889) | 0.920<br>(0.824/0.934) | 0.928<br>(0.927/0.929) | 0.831<br>(0.779/0.915) |
| External set | - | - | - | - | - | - | 0.906<br>(0.835/0.954) |

|  |  |  |  |  |  |  |  |
| --- | --- | --- | --- | --- | --- | --- | --- |
| 3d_fullres default | 0.589<br>(0.509/<br>0.683) | 0.479<br>(0.111/<br>0.573) | 0.858<br>(0.821/0.910) | 0.616<br>(0.402/0.942) | 0.118<br>(0.059/0.181) | 0.619<br>(0.619/0.619) | 0.628<br>(0.553/0.698) |
| 2d default | 0.583<br>(0.494/<br>0.692) | 0.484<br>(0.138/<br>0.668) | 0.819<br>(0.777/0.842) | 0.576<br>(0.533/0.899) | 0.466<br>(0.333/0.600) | 0.632<br>(0.632/0.632) | 0.662<br>(0.492/0.705) |
| Rater 2 | 0.492<br>(0.411/<br>0.614) | 0.720<br>(0.546/<br>0.850) | 0.745<br>(0.627/0.773) | 0.790<br>(0.585/0.832) | 0.792<br>(0.651/0.879) | 0.836<br>(0.832/0.840) | 0.578<br>(0.446/0.646) |
| External set | - | - | - | - | - | - | 0.765<br>(0.684/0.830) |

The checkpoints of the segmentation models can be found in the following github repository: https://github.com/claim-berlin/aSAH-multiclass-segmentation

The experimentation we performed with additional hyperparameter configurations and another in-house model architecture coined BRAVENET (30) can be found in the Appendix (Table 7, Table 8). Overall the nnUnet configured model outperformed the BRAVENET architecture. For reporting our results in the following sections we only focused on the 2d and 3d nnUnet models because no significant performance gains were observed by changing the architecture or tuning its hyperparameters.

Figures 2 and 3 show a few examples of the segmentations of our model on different hemorrhage classes in comparison to the ground truth (Rater 1). As shown the nnUnet is able to detect hemorrhage but struggles to assign the individual voxels to the different hemorrhage classes. The failure cases of the nnUnet models are shown in Figure 6.

**Figure 2:**
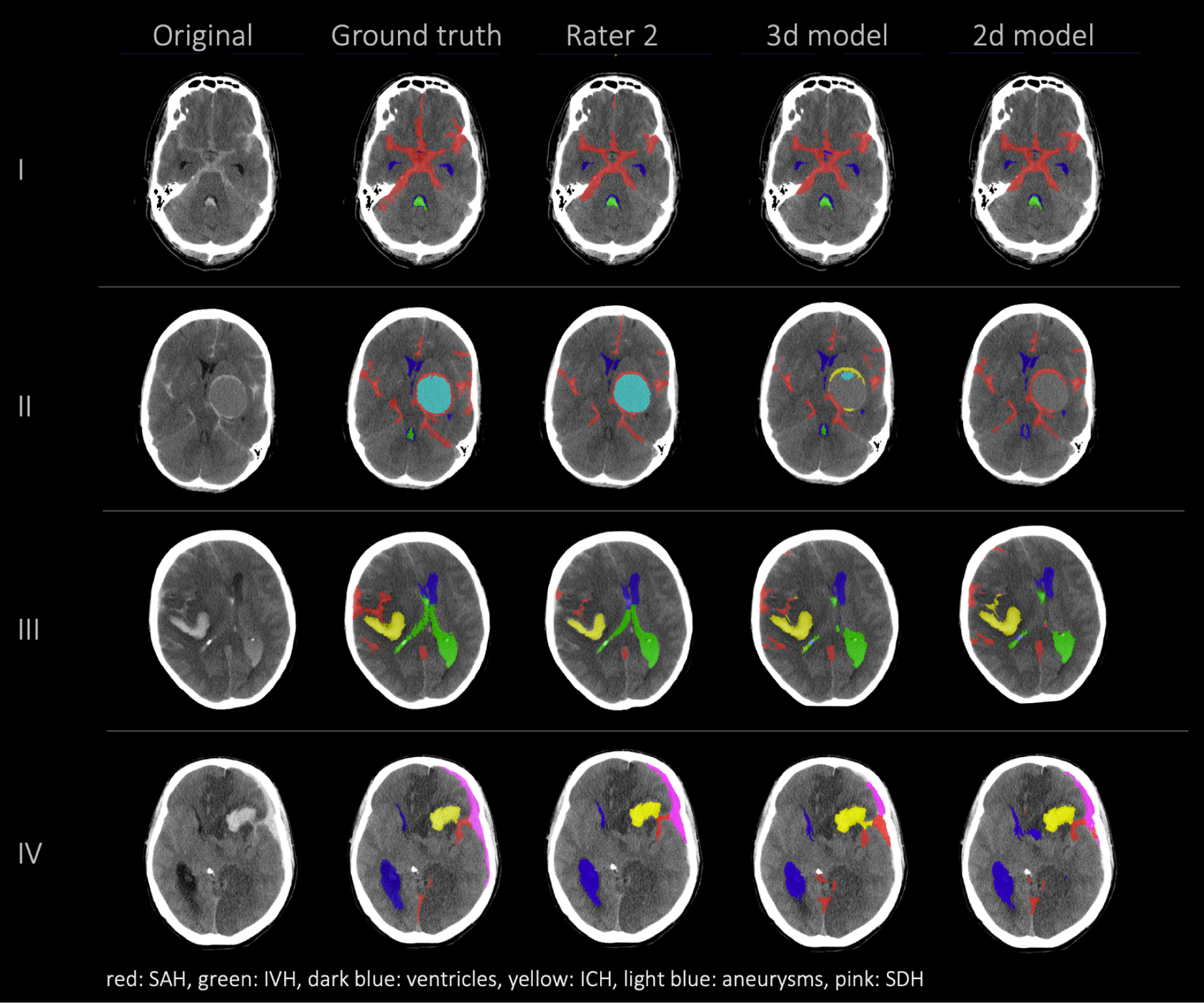
comparison of 4 patients’ segmentations in the following order from left to right: first column shows the original NCCT, second column shows the ground truth (segmentation of Rater 1), third column shows the segmentation of Rater 2 and the fourth and fifth columns show the results of our nnUnet models (fourth column: 3d model, fifth column: 2d model). This figure shows that the nnUnet is able to detect hemorrhage but struggles to assign the segmentations to the different classes. The second row (II) effectively illustrates our findings that in the aneurysm class the results of the nnUnet models and Rater 2 differed the most.

**Figure 3:**
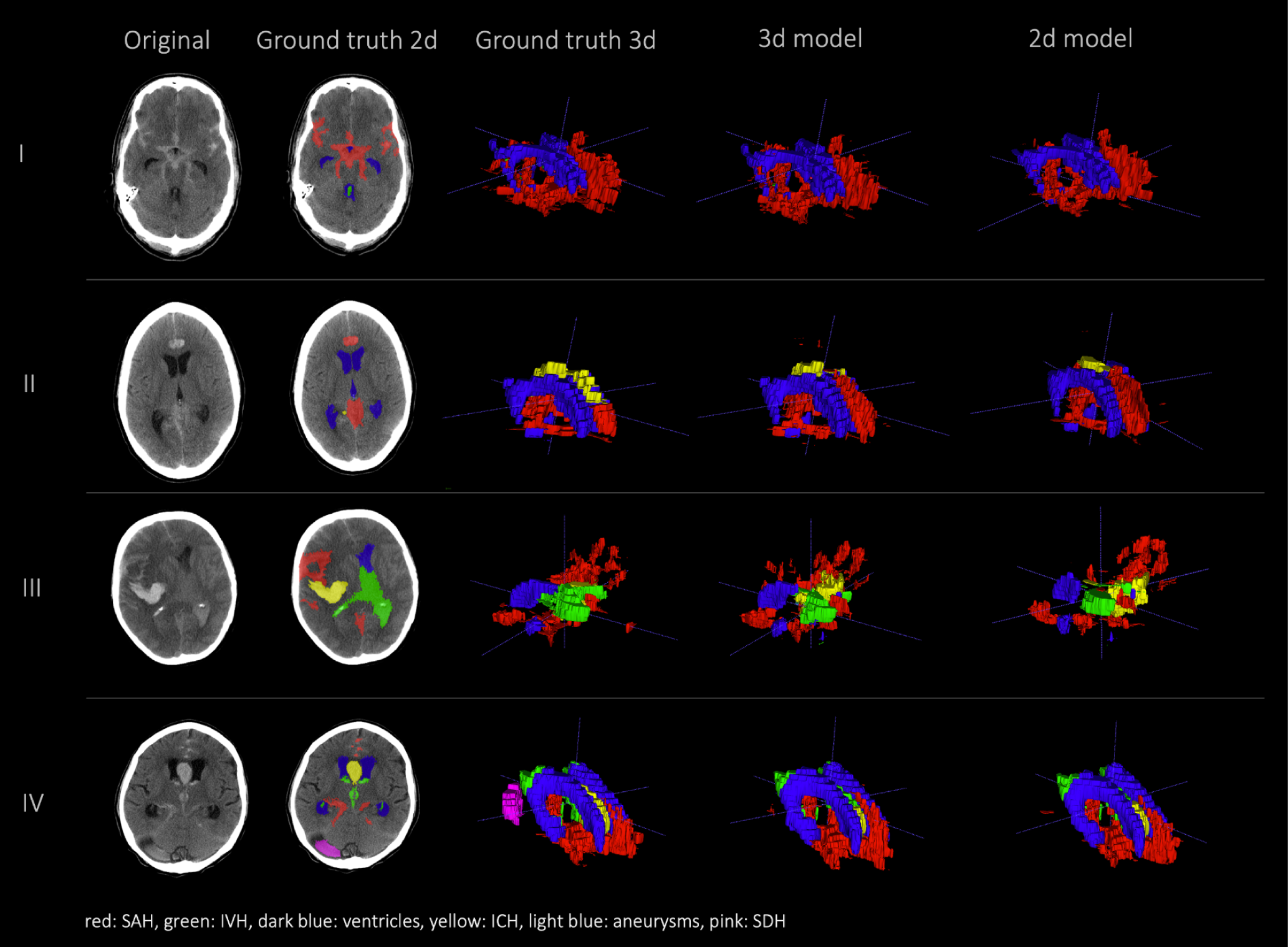
comparison of segmentations of 4 patients in the following order from left to right: first column shows original NCCT, second column shows ground truth (Rater 1 segmentation) in 2d, third column shows 3d representation of ground truth (Rater 1), fourth column shows 3d representation of the 3d model and fifth column shows 3d representation of the 2d model. This figure shows a 3d reconstruction of the hemorrhage distribution which might be useful in clinical practice to get an overview of the extent of the hemorrhage.

### External data set results

In the external dataset the median age was 71 years [min;max = 36;98 years] and 46% of the patients were female. The external dataset included mainly patients with ICH as primary diagnosis, but many patients had co-occurring pathologies like SAH, IVH, and SDH. The automated segmentation of the external data set achieved a global Dice coefficient of 0.838 for the merged hemorrhage class (Table 4 and Figure 5).

**Figure 4.**
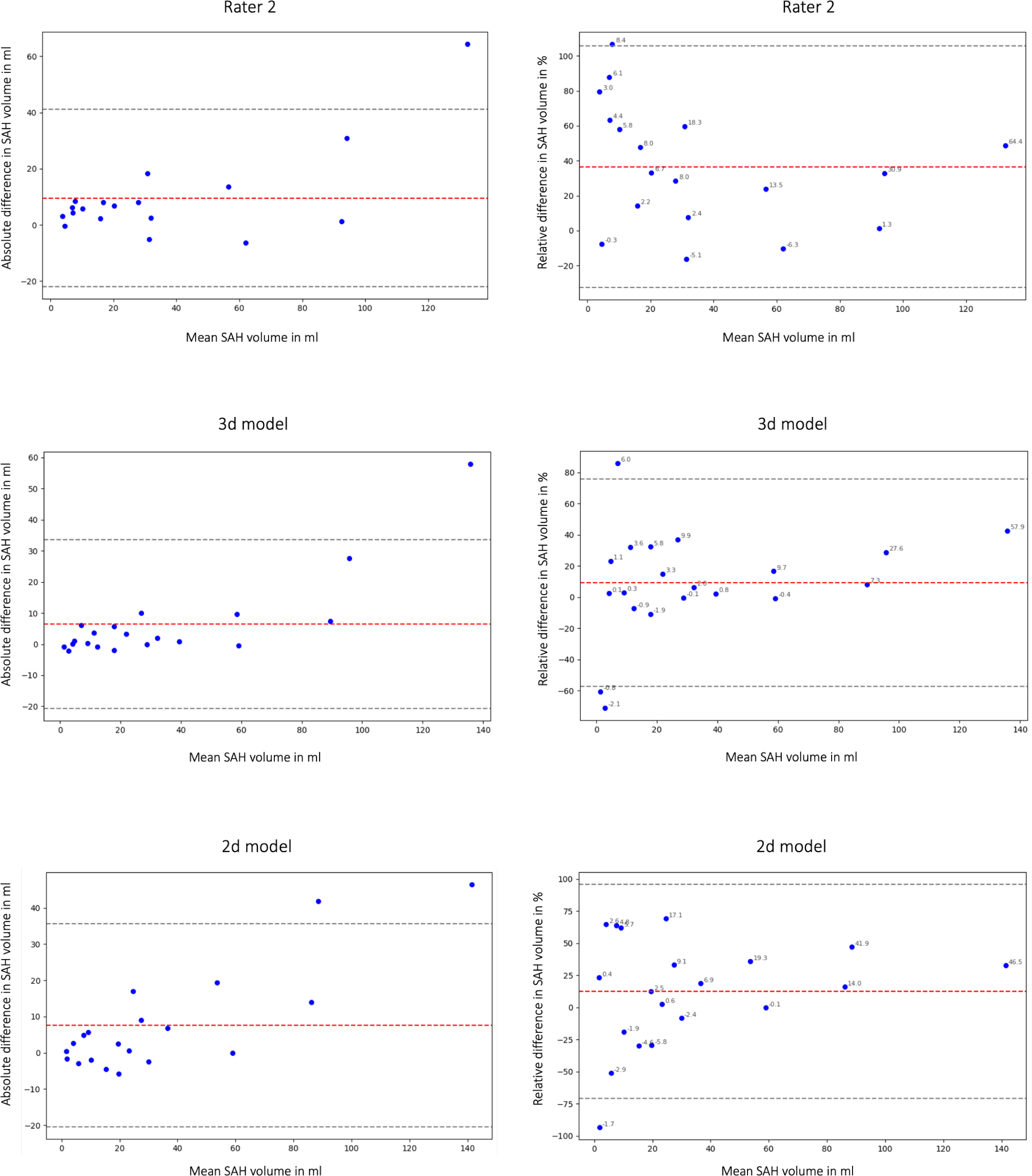
a: Bland Altman Plot absolute (first column) and relative (second column) difference in segmented subarachnoid hemorrhage (SAH) volume. Comparison of segmented subarachnoid hemorrhage volume in ml and % of Rater 2, 3d model and 2d model. Absolute difference in SAH volume: total bias Rater 2: 9.54 ml; total bias 3d model: 6.45 ml; total bias 2d model: 7.59 ml. The average total bias for SAH volume is 7.86 ml which means that Rater 1 (ground truth) segmented 7.86 ml more SAH volume on average than Rater 2, the 3d model and the 2d model. This phenomenon is also shown in Figure 2 and 6.

**Figure 5:**
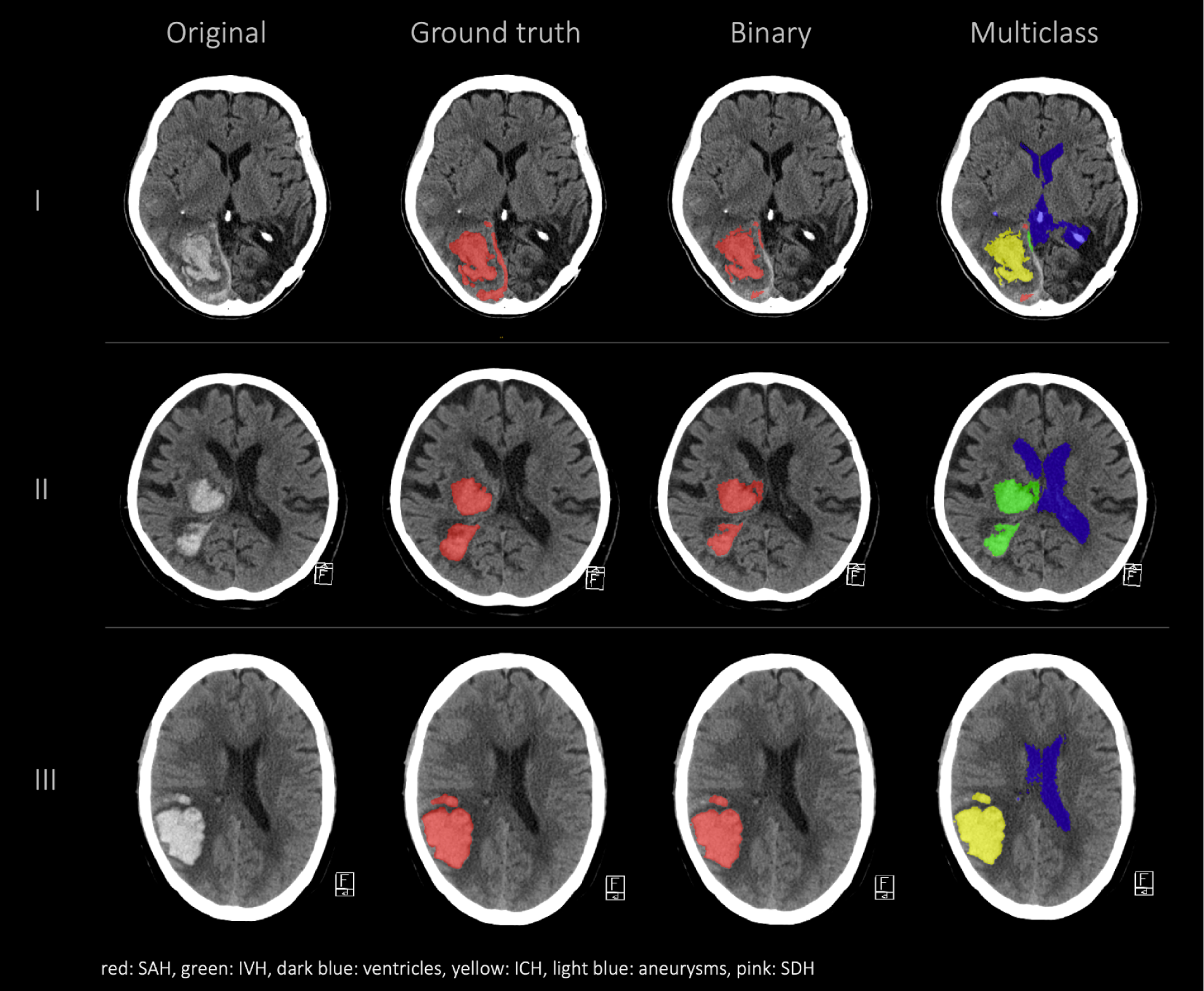
External validation comparison segmentations in the following order from left to right: first column shows the original data, the second column shows the hemorrhage segmentation that was done at an external institution by a human rater (ground truth), the third column shows our 3d model binary segmentation and the fourth column shows our 3d model multiclass segmentation.

**Figure 6:**
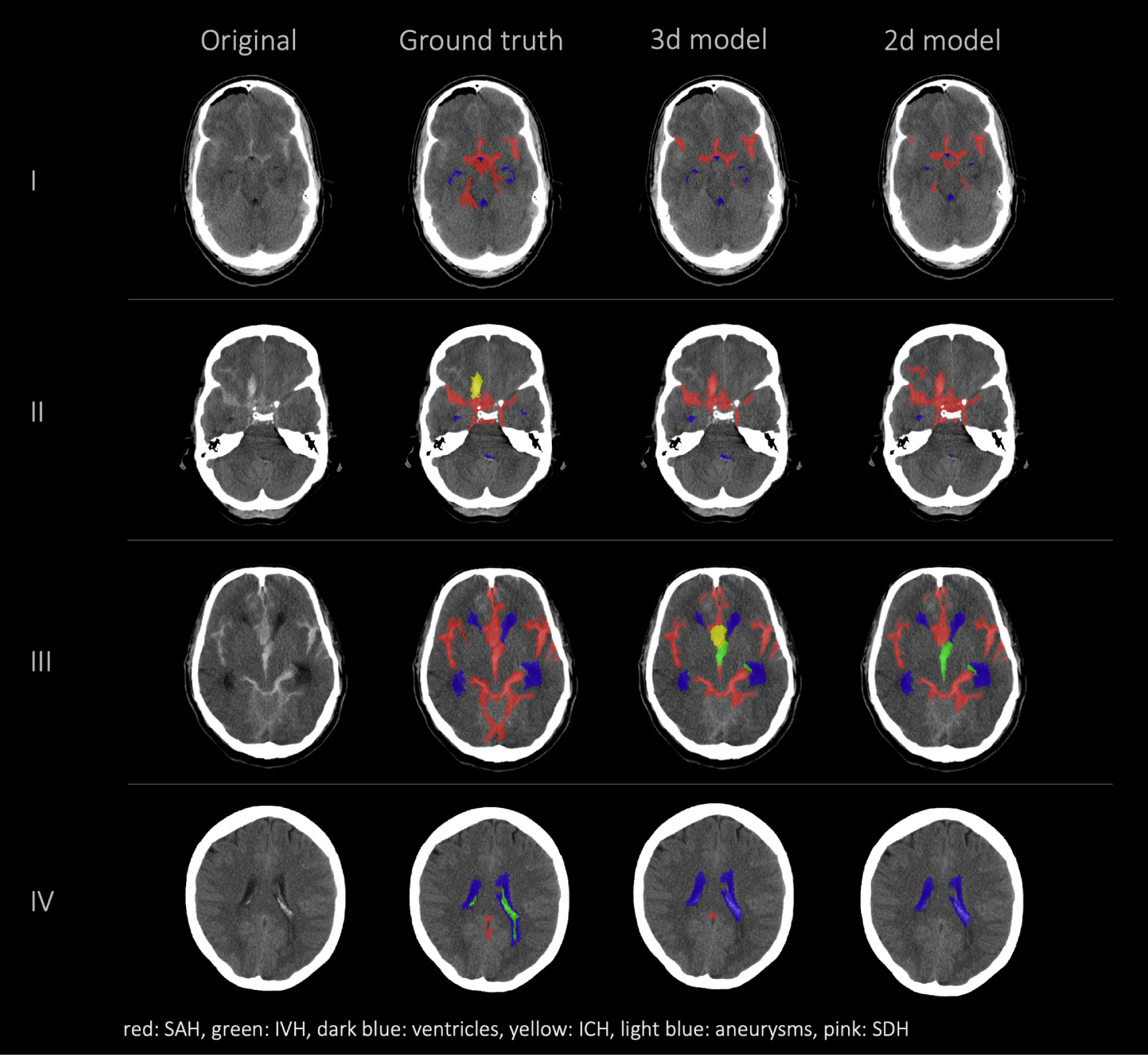
Failure cases, segmentations from left to right: first column shows original data, second column shows the ground truth (segmentation of Rater 1), third and fourth columns show the segmentations of our nnUnet models (segmentation 3d model/segmentation 2d model). This figure showcases the most representative instances of failure we encountered. Once more, it underscores that the challenge does not primarily reside in detecting hemorrhage but rather in accurately distinguishing between various types of hemorrhages.

As shown in Table 5 both of the nnUnet models agreed with the ground truth of Rater 1 on the presence of SAH in all cases, whereas Rater 1 and Rater 2 did not agree on the presence of SAH in two patients. The two different nnUnet models agreed with the ground truth on the presence of IVH, ICH, and SDH in 8/5/1 cases. The two nnUnet models disagreed with the ground truth mostly on the presence of IVH and ICH (3d model: 3/4; 2d model: 4/4), whereas Rater 2 disagreed on a relatively similar number of cases in the IVH, ICH, aneurysm and SDH class (2/3/2/1). The ventricle class showed no disagreements between the ground truth, the 3d model, the 2d model, and Rater 2. The agreements and disagreements for all classes are shown in Table 5.

**Table 5:** Agreement/Disagreement 3d model/2d model/Rater 2. The first column illustrates the agreements in labeling the different classes between the ground truth (Rater 1) and the 3d model (first row), 2d model (second row), Rater 2 (third row). The second column shows the disagreements in labeling the classes and the third column shows the number of NCCT where a certain class was not present, therefore we can not assess the agreement/disagreement.

|  | Agreement |  | Disagreement |  | Class not present |  |
| --- | --- | --- | --- | --- | --- | --- |
|  | SAH | IVH//ICH//Aneurysm//SDH | SAH | IVH//ICH//Aneurysm//SDH | SAH | IVH//ICH//Aneurysm//SDH |
| 3d_fullres default | 20 | 8//5//3//1 | 0 | 3//4//1//1 | 0 | 9//11//16//18 |
| 2d default | 20 | 8//5//2//1 | 0 | 4//4//2//1 | 0 | 8//11//16//18 |
| Rater 2 | 18 | 9//4//3//2 | 2 | 2//3//2//1 | 0 | 9//13//15//17 |

In this study we not only assessed the Dice coefficient but also calculated the volumetric similarity (VS) to create a connection between the correctly segmented volume and the overall hemorrhage volume. Figure 4a shows the volumetric similarity for the SAH class using Bland Altman Plots and therefore the absolute and relative difference between the ground truth, the segmented SAH volumes by Rater 2 and by the 3d and 2d nnUnet model. It illustrates that even though smaller volumes have smaller differences in segmented volume (Figure 4a: left column) relatively speaking, the deviation based on the total segmented volume is larger (Figure 4a: right column).

## Discussion

We present in the current work a deep learning based analysis of automated multiclass segmentation for SAH-related pathologies from NCCTs of patients with SAH. Our framework demonstrates a high quantitative performance based on global Dice coefficient and volumetric similarity that is close to the performance of a human rater. We show that our nnUnet models are able to do a precise and fast segmentation of aSAH related classes that are relevant to assess patient outcome.

### Our results

We evaluated our model with quantitative measures, compared performance against interrater variability of medical expert raters, and performed an external validation. Overall, despite a limited training dataset, we attain highly promising results in most classes and demonstrate generalizability on an external dataset featuring ICH as primary hemorrhage class.

We observe varied performance across the different classes and identify important findings that might explain these differences in performance. Overall, the two deep learning models, 3d model and 2d model, show a similar performance. Segmentations of SAH, ventricles, ICH and the hemorrhage segmentation are comparable in 3d model, 2d model and Rater 2 whereas the aneurysm and SDH class show the highest variability. The difference between the performance of the nnUnet models in comparison to Rater 2 is relatively high in the aneurysm and SDH class. This is most likely due to the limited occurrence of these classes in our training set and the limited size of our internal test set. Compared to the segmentation of the SAH class, better results were achieved for the IVH, ICH and SDH class by the nnUnet models. The interrater reliability was assessed to create human benchmarks which can be used to put our models’ results into context. A comparison of the models’ results to human benchmarks is necessary because absolute values of metrics are challenging to interpret and thus model performance can be over-/underestimated. For instance, from an absolute value point of view, a global Dice coefficient of 0.686 for SAH is lower than a global Dice coefficient of 0.758 (3d model)/0.766 (2d model) for SDH but actually compared to human benchmarks (Rater 2 global Dice coefficient of 0.665 for SAH and 0.902 for SDH) the overall automated segmentation performance for SAH is closer to the human performance and is considered a better result when aiming for human-like performance. In this sense, optimizing for higher performance - with an improvement of the global Dice coefficient - with architectural changes or optimizing the training approach is not the main goal of our study.

Interestingly, we achieve a superior performance according to the global Dice coefficient in the external test dataset compared to the internal test set for the ICH class. This is a relatively uncommon occurrence in the field of machine learning and is most likely due to the presence of SAH in our internal dataset making the labeling of the ICH more challenging. This phenomenon was also reported in the INSTANCE Grand challenge where SAH achieved the worst results compared to all the other intracranial hemorrhages that were evaluated (SDH, epidural hematoma (EDH), IPH, IVH) (20). Wu et al. also reported the best results for multiclass segmentation for IVH and ICH and an inferior performance for SAH, SDH and EDH (25). This highlights the challenging nature of distinguishing between different hemorrhage subtypes in real world cases of SAH.

Hemorrhage segmentation can also become more difficult in patients with shunts and metal artifacts (Appendix: Figure 7) due to previous bleeding or intensive-care-unit (ICU)-monitoring. Shunt artifacts could be misinterpreted as ventricles and metal artifacts might resemble SAH (Appendix: Figure 7). Being aware of potentially inferior performance in scans with artifacts, we include those patients to improve model generalizability and robustness in cases where a metal artifact or shunt was present.

The ventricle class achieves the best results out of all classes most likely because of its consistent shape and location and because it is the most represented class in the dataset. Nonetheless it is important to consider the size and shape of ventricles in SAH patients because they can be altered due to increased intracranial pressure and provide valuable information concerning the patients outcome e.g. enlarged ventricles can be a sign of increased intracranial pressure which can lead to decrease of cerebral perfusion pressure and therefore an unfavorable outcome (23). Ventricles as one of the most significant anatomical structures in SAH can serve as an anatomical orientation for downstream outcome prediction models. Therefore the segmentation of the ventricles is relevant and can be considered an advantage of our model compared to already published automated multiclass segmentation models (20,25).

### 2d/3d model

The 2d and 3d model architectures performed similarly based on quantitative assessments by global Dice coefficient and volumetric similarity. However, there were some locations (Figure 2: aneurysm, SDH) where it is difficult for the model to distinguish the type of the bleeding looking only at one axial slice.

The use of 2d models can be more advantageous for generalization to volumes with various slice thicknesses because 3d models either require reslicing or might show poor performance when the voxel spacings are significantly different on the test sets. However, as stated in the INSTANCE Grand challenge, directly utilizing 2d networks would lead to a loss of significant context information among slices (20). Additionally the spatial distribution of SAH in three dimensions is not considered in most of the known radiographic scores but can be important for outcome prediction.

### Current use of deep learning-based segmentation

Radiological features of hemorrhages in aSAH patients provide valuable outcome relevant information that is not exploited to date. Despite the general acceptance that the volume of blood after SAH is prognostic for outcome and provides guidance for treatment decisions, no method to estimate the volume and distribution patterns of subarachnoid blood has been successfully implemented in clinical practice so far.

As stated in a recent article about AI in medicine and clinical practice there is a need to not only assess the stand-alone performance of models but more so focus on the outcomes when these algorithms are used as assistive tools in clinical practice (32). In our study, achieving performance levels comparable to human assessment based on quantitative metrics is feasible across most classes, despite the constraints of limited data. However, what is interesting for future investigations is assessing the model’s efficacy when utilizing segmentation outputs in a downstream task aimed at predicting outcomes in patients with SAH. There are a few examples for radiomics-based classification of intracranial pathologies e.g., intracranial aneurysm rupture or SAH prognosis prediction (33–35). In future works, the extraction of radiomics features based on segmentation masks might allow the extraction of imaging biomarkers predictive of SAH outcome. Deep learning based segmentation models presented in this study would not only allow to distinguish between different types of hemorrhages and healthy brain tissue, but also give precise information about the exact volumes of different structures and the impact on the patient’s outcome.

There are a few examples of automated segmentation in NCCT in patients with SAH or ICH as leading pathology (16,20,36). The different models achieved Dice scores that were close to human level performance via volume quantification for SAH based on density quantification (16) or intracerebral hemorrhage segmentation via viola-Unet (“Voxels-Intersecting Along Orthogonal Levels Attention U-Net”) (36) in the INSTANCE Grand Challenge 2022. Even though the studies included patients with different subtypes of intracranial bleeding, only a recent multiclass model from Wu et al. was able to distinguish between different subtypes of intracranial hemorrhage (25). However, since anatomical changes, e.g. altered shape of ventricles, can be a negative predictor for the patient’s outcome, it is important to assess the shape and volume of the ventricles as well and not only report the different types of hemorrhages.

Co-occurring pathologies can change the outcome of SAH-patients drastically for the worse, creating the need for a tool that is able to distinguish between the different subtypes of hemorrhage and can segment anatomical structures like ventricles (23,24). Automated multiclass segmentation in NCCT was only tested on patients with ICH and intracranial hemorrhage in general so far, not specifically for patients with SAH (25,37,38).

### Study limitations

Our study had several limitations. First, the number of patients in our dataset was limited due to the time-consuming manual segmentation step and resulted in the underrepresentation of some classes in the training set as well as in the validation and test set (e.g. aneurysm, SDH), which could lead to an inferior performance of the models for these classes. Additionally, to validate the performances of the nnUnet models, further training with a larger dataset may be beneficial. Second, the model is trained mostly on patients with subarachnoid hemorrhage to quantify the volume and distribution of hemorrhage subtypes. Hence, the models are not directly applicable for classifying CTs whether they contain hemorrhage or healthy brain tissue.

Additionally, our external validation set does not include primarily SAH patients. Despite being tested on a different patient population our model demonstrated considerable generalization. Future works should test the performance of our model in segmentation challenges and public multiclass cerebral hemorrhage segmentation datasets.

In this study we aim to compare our models’ performance to human benchmarks which we created by assessing the performance of a different human rater (Rater 2) other than the ground truth (Rater 1). The comparison to human level performance is likely more relevant in the clinical context but needs presumably more than 2 different human Raters and a consensus based ground truth creation to validate the comparison of the performances of humans and our nnUnet models.

### Future Work

Our work has implications for future works in SAH segmentation and outcome prediction. Traditional scores lack interrater and intrarater reliability and often create disagreements between different reviewers. All known radiographic scores calculate the amount of blood via hemorrhage thickness estimations in different areas in NCCT which is either roughly precise (modified Fisher scale, Hijdra Scale, (7,9)) or only applicable for symmetrical hemorrhages (ABC/2 Scale (39)), which leads to a low interrater reliability. A high interrater reliability ensures consistency in the segmentations irrespective of different rater, it provides a measure of agreement among human raters and is serving as a benchmark for evaluating the performance of automated methods. In clinical context, it ensures quality control, reproducibility and reduces bias. Because in clinical context the exact determination of the hemorrhage volume and severity of the hemorrhage is crucial for the treatment decision and outcome of the patient (23), there is a strong clinical need for a score or model that is reliable and shows consistent performance regardless of qualification or experience of the clinical personnel.

The prognostic value of different traditional radiographic scores can be increased by an automated calculation that is supported by deep learning methods like our model or other radiomics approaches (35,40). Additionally, the multiclass nature of our proposed model can be utilized in future studies to analyze the correlation of outcomes with co-occurrence of hemorrhage classes, their spatial distribution, hemorrhage volume and radiomics signatures. Our framework can be used to segment a large number of patients, enabling the processing of extensive patient data while being as precise as manual segmentation without being time-consuming.

With the help of deep learning there is the possibility to develop novel scores and biomarkers based on the exact imaging information. Deep learning-based automated semantic segmentation can facilitate the extraction of volumetric information from CT scans and enable an objective assessment of SAH severity. Development of novel prognostic approaches that overcome limitations of traditional scores - pending further validation - can enable rater-independent outcome prediction. These approaches can result in the extraction and utilization of more radiographic data from CT scans, providing additional information that enables outcome prediction and facilitates decision support for personalized treatment decisions to improve patient outcomes.

## Conclusion

Our deep learning-based nnUnet-model demonstrated a performance close to the human benchmark and achieved accurate segmentation of SAH and SAH-related pathologies. This can be the starting point for automation of traditional radiographic scores, correlation analysis of outcomes with co-occurence of hemorrhage classes and development of novel, prognostic scores for predicting outcomes in SAH. Deep learning can overcome the significant limitations of interrater and intrarater variability and provide an efficient solution to better exploit outcome-relevant image information than traditional scores. Our open source model can enable analyses of large multicentric datasets to further improve performance and explore generalization.

Taken together, our study demonstrates the potential of deep learning to improve patient outcomes by advancing radiographic examination of SAH and other intracranial hemorrhages.

## Data Availability

All data produced in the present study are available upon reasonable request to the authors. Our model is made open source with pre-trained weights to facilitate the extraction of outcome-related pathologies from NCCT images of aSAH patients for further research.

https://github.com/claim-berlin/aSAH-multiclass-segmentation

## Abbreviations

SAH: subarachnoid hemorrhage
aSAH: aneurysmal SAH
ICH: intracerebral hemorrhage
IVH: intraventricular hemorrhage
SDH: subdural hematoma
NCCT: non-contrast CT scans
CNN: convolutional neural network
BNI: Barrow Neurological Institute
DICOM: Digital imaging and communications in medicine
NIfTI: Neuroimaging informatics technology initiative
TP: true positives
FP: false positives
FN: false negatives
DCI: delayed cerebral ischemia
SEBES: subarachnoid hemorrhage early brain edema score
VS: volumetric similarity
EDH: epidural hematoma
IPH: intraparenchymal hemorrhage
ICU: intensive care unit

## Acknowledgements

Computation has been performed on the HPC for Research cluster of the Berlin Institute of Health.

