## Appendix for "Deep Learning-based Multiclass Segmentation in Aneurysmal Subarachnoid Hemorrhage"

### Appendix I

| | SAH | IVH | Ventricles | ICH | Aneurysm | SDH | hemorrhage class |
|---|---|---|---|---|---|---|---|
| **mean Dice ± STD** | | | | | | | |
| 3d_fullres default | 0.597 ± 0.126 | 0.474 ± 0.312 | 0.850 ± 0.056 | 0.665 ± 0.140 | 0.183 ± 0.172 | 0.758 ± n.d. | 0.633 ± 0.132 |
| 2d default | 0.587 ± 0.140 | 0.485 ± 0.315 | 0.848 ± 0.053 | 0.698 ± 0.176 | 0.541 ± 0.298 | 0.766 ± n.d. | 0.623 ± 0.157 |
| Rater 2 | 0.576 ± 0.143 | 0.680 ± 0.276 | 0.810 ± 0.071 | 0.652 ± 0.401 | 0.790 ± 0.125 | 0.896 ± 0.013 | 0.630 ± 0.170 |
| External set | - | - | - | - | - | - | 0.776 ± 0.155 |
| **mean volumetric similarity ± STD** | | | | | | | |
| 3d_fullres default | 0.879 ± 0.124 | 0.649 ± 0.332 | 0.943 ± 0.046 | 0.769 ± 0.188 | 0.278 ± 0.300 | 0.775 ± n.d. | 0.865 ± 0.117 |
| 2d default | 0.822 ± 0.126 | 0.671 ± 0.268 | 0.927 ± 0.049 | 0.818 ± 0.189 | 0.660 ± 0.447 | 0.786 ± n.d. | 0.826 ± 0.125 |
| Rater 2 | 0.798 ± 0.153 | 0.737 ± 0.273 | 0.881 ± 0.075 | 0.680 ± 0.415 | 0.865 ± 0.119 | 0.928 ± 0.002 | 0.810 ± 0.147 |
| External set | - | - | - | - | - | - | 0.870 ± 0.132 |
| **mean sensitivity ± STD** | | | | | | | |
| 3d_fullres default | 0.584 ± 0.140 | 0.412 ± 0.315 | 0.852 ± 0.081 | 0.664 ± 0.289 | 0.120 ± 0.122 | 0.619 ± n.d. | 0.604 ± 0.126 |
| 2d default | 0.573 ± 0.159 | 0.431 ± 0.312 | 0.812 ± 0.086 | 0.654 ± 0.237 | 0.466 ± 0.378 | 0.632 ± n.d. | 0.589 ± 0.162 |
| Rater 2 | 0.503 ± 0.160 | 0.615 ± 0.303 | 0.730 ± 0.107 | 0.628 ± 0.404 | 0.756 ± 0.230 | 0.836 ± 0.011 | 0.545 ± 0.174 |
| External set | - | - | - | - | - | - | 0.725 ± 0.160 |

Table 6: summary of the results of the mean values of our models, Rater 2 and the external set.

| | SAH | IVH | Ventricles | ICH | Aneurysm | SDH | hemorrhage class |
|---|---|---|---|---|---|---|---|
| **median Dice** | | | | | | | |
| 3d_fullres default | 0.5805 | 0.5679 | 0.8622 | 0.6218 | 0.2082 | 0.7584 | 0.6636 |
| 2d default | 0.6157 | 0.5146 | 0.8551 | 0.6920 | 0.5406 | 0.7664 | 0.6725 |
| BRAVENET_version_1 | 0.5418 | 0.2933 | 0.8401 | 0.5587 | 0.2142 | 0.3374 | 0.6286 |

| | | | | | | | |
|---|---|---|---|---|---|---|---|
| BRAVENET_version_3 | 0.5689 | 0.4925 | 0.8547 | 0.7175 | 0.0394 | 0.6557 | 0.6496 |
| Rater 2 | 0.6263 | 0.7919 | 0.8084 | 0.8339 | 0.8332 | 0.8961 | 0.6593 |

Table 7: comparison of the median Dice results including both nnUnet models (3d_fullres default, 2d default), two examples of the BRAVENET models (BraveNet_version_1, BraveNet_version_3) and Rater 2. The BRAVENET models were integrated into the nnUnet framework for training and interference. We are referring to the BRAVENET repository for further details.
https://github.com/hilbysfe/brain-vessel-segmentation

### Appendix II

| | BraveNet_version_1 | BraveNet_version_2 | BraveNet_version_3 |
|---|---|---|---|
| Param. Num. | 15,379,925 | 66,164,309 | 139,664,618 |
| Stages Num. | 4 | 4 | 7 |
| Kernel Size | 3x3x3 | 5x5x5 | 3x3x3 |
| Base Filters Num. | 32 | 32 | 32 |
| Max Filters Num. | 320 | 320 | 320 |
| Patch Size | 256x256x8 | 256x256x8 | 256x256x8 |
| Context Size* | 512x512x16 | 512x512x16 | 512x512x16 |
| Activation Function | ReLU | ReLU | ReLU |
| Dropout | 0.1 | 0.1 | 0.1 |
| Norm. Op. | InstanceNorm (eps = 1e-5) | InstanceNorm (eps = 1e-5) | InstanceNorm (eps = 1e-5) |

Table 8: Hyperparameters of the BRAVENET models. *The context data is created by extracting a slice from the original data, centered around the bounding box with dimensions extended to ensure the final context size is twice the patch size. In instances where the context exceeds the data bounds, padding is added accordingly.

### Appendix III

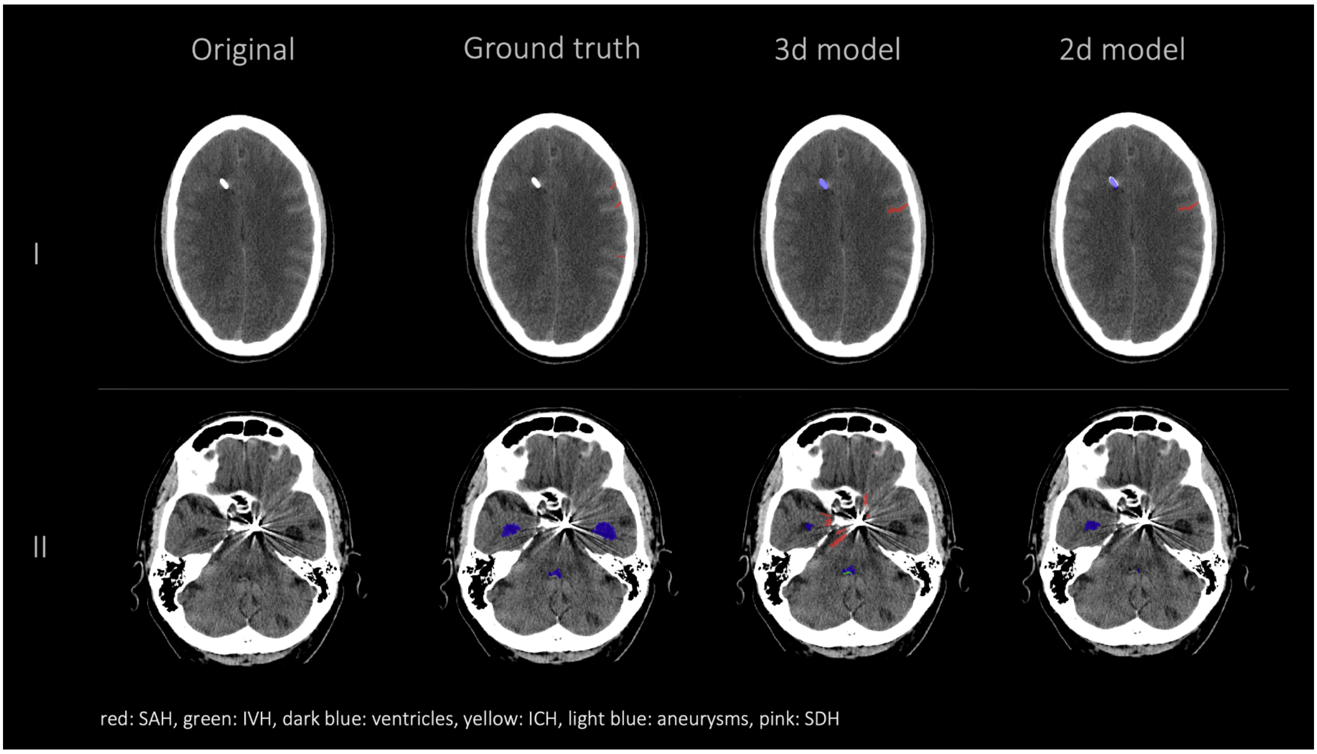


Figure 7: Examples for failure classes with shunt and metal artifacts. First column shows the original data, the second column shows the ground truth (Rater 1), the third column shows the segmentation of our 3d model and the fourth column shows the segmentation of our 2d model. As illustrated artifacts complicate the segmentations of nnUnet models. The shunt can be mistaken for ventricles (as shown in row I) or hemorrhage. Metal artifacts degrade the quality of the NCCT making it challenging to segment structures that are obscured by the artifact.

### Appendix IV

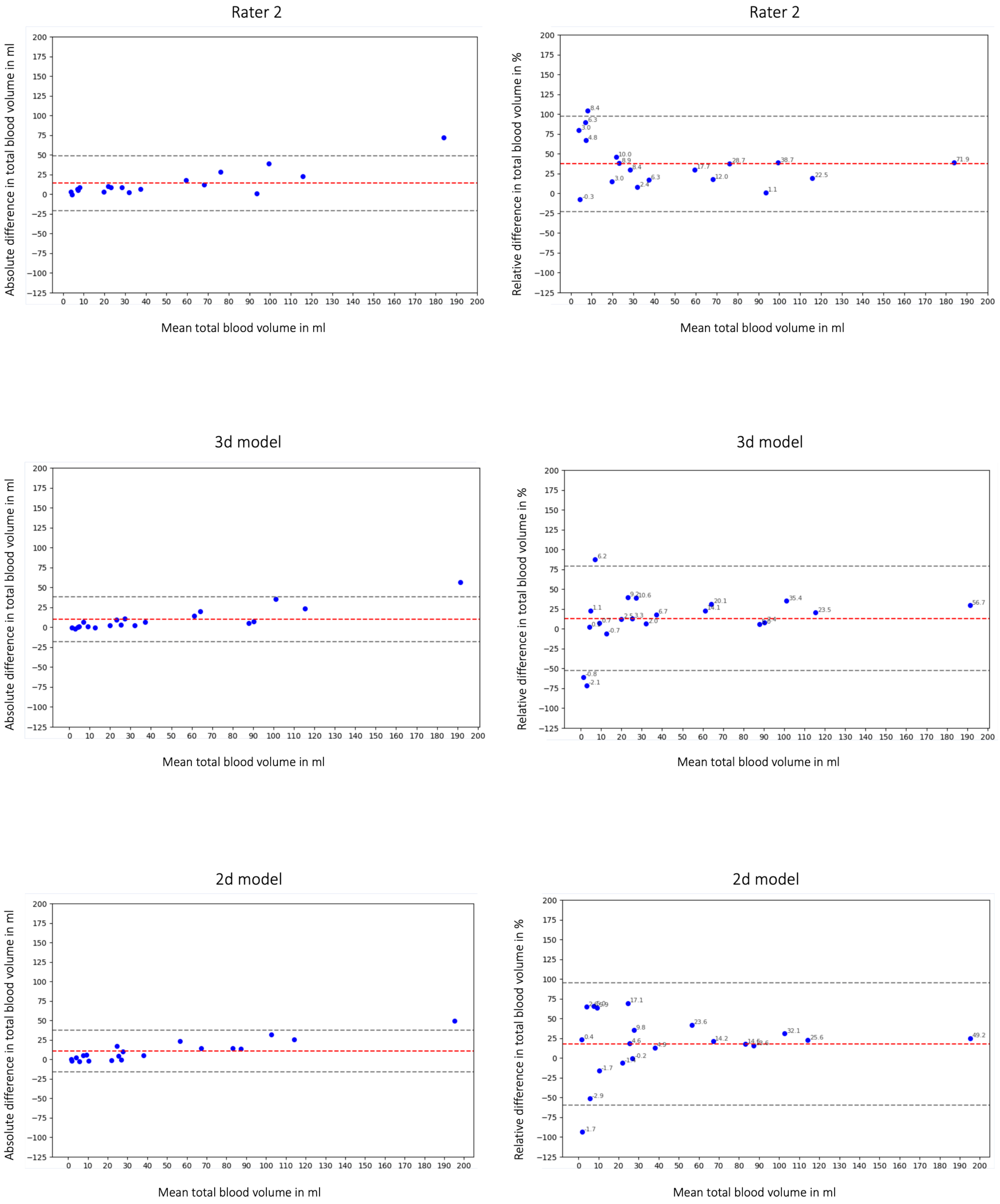


Figure 4b: Bland Altman Plot absolute (first column) and relative (second column) difference in segmented total hemorrhage volume. First row shows Rater 2, second row the 3d model and third row the 2d model. Absolute difference in total

hemorrhage volume: total bias Rater 2: 14.10 ml; total bias 3d model: 10.05 ml; total bias 2d model: 10.76 ml. The average total bias is 11.64 ml which means that Rater 1 (ground truth) segmented on average 11.64 ml more hemorrhage than Rater 2, the 3d model and the 2d model. This phenomenon is also shown in Figure 2 and 6.
